# Psychometric Properties of a 20-Item Adapted Version of the Ontario Child Health Study Emotional Behavioural Scales for Population-Based Research

**DOI:** 10.64898/2026.09.26.26364070

**Authors:** Emma Nolan, Li Wang, Stelios Georgiades, Peter Szatmari, Laura Duncan, Harriet MacMillian, Katholiki Georgiades

**Author notes:** **Corresponding Author:** Dr Emma Nolan.

## Abstract

**Background:** The 52-item Ontario Child Health Study Emotional Behavioural Scales (OCHS-EBS) has demonstrated reliability and validity evidence in Canadian population-based samples. This study evaluated an adapted item set of the Ontario Child Health Study Emotional Behavioural Scales (OCHS-EBS-A) intended to measure selected mental disorder symptoms while reducing respondent burden in population-based research.

**Methods:** A prespecified candidate set of 21 items from selected OCHS-EBS disorder scales assessed depression (4 items), generalized anxiety (4 items), oppositional behaviour (6 items), and attention-deficit/hyperactivity (7 items). The 2023 Longitudinal Canadian Health Survey on Children and Youth (L-CHSCY; ages 5–21 years; n= 22,740) provided calibration and internal-validation samples. The Ontario Provincial Assessment and Tracking of Child and Family Health Study (ONPATH; Wave 5: n=719 parent and n=116 youth respondents) provided an independent sample for further evaluation. Analyses examined factorial validity, internal consistency reliability, convergent and discriminant validity, and measurement invariance across sex assigned at birth within age/informant groups.

**Findings:** The analyses supported a refined 20-item OCHS-EBS-A with three correlated factors representing internalizing, oppositional behaviour, and inattention/hyperactivity symptoms. Prespecified criteria for internal consistency reliability and convergent and discriminant validity were met within the reported L-CHSCY age/informant groups. Measurement invariance across sex assigned at birth was supported in the L-CHSCY groups and for children and parents in and youth-reported internalizing symptoms in ONPATH. However, not for ONPATH youth-reported oppositional behaviour symptoms.

**Interpretation:** Results from these population-based samples provide factorial-validity, internal-consistency, and convergent- and discriminant-validity evidence for scores from the 20-item OCHS-EBS-A among children, youth, and emerging adults aged 5–21 years.

## Introduction

Measurement of child and youth mental health in the general population is essential for informing mental health policy, prevention, and intervention strategies. Brief measures of child and youth mental disorder symptoms are important for population-based research because structured diagnostic interviews are costly and time-consuming at scale (Duncan, Wang, & Boyle, 2024). Dimensional symptom-based measures provide a feasible alternative, but their utility depends on their psychometric properties, and evidence that they function consistently across developmental stages, informants, and population contexts. Despite their widespread use, relatively few brief measures have undergone comprehensive psychometric evaluation in large population-based samples (Achenbach, 2011; Kieling et al., 2024).

The original 52-item Ontario Child Health Study Emotional Behavioural Scales (OCHS-EBS) is a multidimensional measure of symptoms aligned with selected DSM-5 disorder domains: conduct disorder (CD), oppositional defiant disorder (ODD), attention-deficit/hyperactivity disorder (ADHD), major depressive disorder (MDD), social anxiety disorder, separation anxiety disorder, and generalized anxiety disorder (Duncan et al., 2019) The OCHS-EBS has a long history of development and use in Canadian epidemiological surveys, with roots in the original 1983 Ontario Child Health Study and refinements over subsequent decades. It is freely available, linked to selected DSM symptom criteria, and can support Canadian population norms. Any adapted item set nevertheless requires evidence that remains reliable and valid across the ages, informants, and survey contexts in which it will be used. Previous evaluation using parent and youth reports in the 2014 Ontario Child Health Study generally supported the proposed structure and score reliability and validity. The CD scale demonstrated comparatively weaker internal consistency among younger children and did not meet all psychometric criteria (Duncan et al., 2019).

Subsequent studies have evaluated the OCHS-EBS in additional samples and formats, including an outpatient sample and a 25-item OCHS-EBS-Brief (OCHS-EBS-B) (Boyle et al., 2022; Duncan et al., 2025; Ferro et al., 2024). The 25-item OCHS-EBS-B was developed using multidimensional Item Response Theory (IRT) methods to identify items with strong discrimination and appropriate difficulty parameters while reducing the number of symptom domains represented (Boyle et al., 2022). Consequently, the brief version did not retain all domains included in the original 52-item OCHS-EBS, instead focusing on a subset of emotional, behavioural, and attentional symptom dimensions most suitable for brief assessment. The brief scale has been evaluated both as a categorical (binary) classification tool and as a dimensional measure. Validation studies have focused on the parent/caregiver report of children aged 4-17 years (Boyle et al., 2022), and demonstrated good reliability and construct validity as both a categorical and dimensional measure in population-based and mental health service samples. Internal consistency exceeded .80 across age groups, test–retest reliability was high (r = .86–.87), and confirmatory factor analyses supported a three-factor structure representing emotional, behavioural, and attentional symptom domains. Subsequent work has provided additional evidence supporting its use in clinical and health service settings (Ferro et al., 2024). Collectively, these findings provide strong psychometric evidence for use of the OCHS-EBS in population-based epidemiologic surveys and clinical samples of children and youth accessing mental health services. These studies provide relevant evidence across parent informants, age groups, and settings.

Less evidence is available regarding the psychometric performance of the OCHS-EBS items in more recent and national-population-based, general health surveys, particularly among emerging adults. National, population-based surveys increasingly rely on brief measures to assess mental health symptoms efficiently and at scale. The utility of such measures depends on their ability to provide reliable, valid, and developmentally appropriate assessments across diverse age groups. The item set examined in the present study is described as adapted rather than abbreviated or brief. The established OCHS-EBS-B was not well aligned with the aims of the data collection and study because its item set includes conduct disorder and specific anxiety disorder domains, whereas we sought to focus on major depressive disorder, generalized anxiety disorder, and oppositional defiant disorder. We therefore selected candidate items from the full 52-item OCHS-EBS, excluding conduct disorder and specific anxiety disorder items and expanding the coverage of oppositional defiant disorder. Given these substantive differences from the established brief form, we refer to this item set as the Ontario Child Health Study Emotional Behavioural Scales– Adapted (OCHS-EBS-A) and evaluate its psychometric performance in a national, population-based sample that includes emerging adults.

The present study extends previous work by evaluating the OCHS-EBS-A in a substantially larger, population-representative sample spanning all 10 Canadian provinces and within the age and informant configurations used in two contemporary national surveys. It also expands the age range examined in the original OCHS-EBS development work to include emerging adults aged 18–21 years. Finally, the study provides evidence from a more recent, post-COVID-19 context, in which concerns about the mental health of young people remain pronounced, and evaluates whether reliability and validity criteria are met within each available age and informant group.

The 2023 Longitudinal Canadian Health Survey on Children and Youth (L-CHSCY), a population-based follow-up survey covering all ten Canadian provinces, was used to develop and internally validate the factor structure. The Ontario Provincial Assessment and Tracking of Child and Family Health Study (ONPATH), an independent Ontario population-based study, was used to evaluate the resulting structure in an independent sample. Evaluation among emerging adults aged 18–21 years is important because this developmental period is associated with the onset and persistence of many mental disorders (Arnett, 2000; Kessler et al., 2005; Kieling et al., 2024). Extending an item set beyond its previously evaluated age range requires explicit evidence for its factorial validity, score reliability, and measurement invariance in that group.

This study evaluated the psychometric properties of the adapted OCHS-EBS item set, including factorial validity, internal consistency reliability, convergent and discriminant validity, and measurement invariance. We hypothesized that a three-factor structure representing internalizing, oppositional behaviour, and attention deficit/hyperactivity symptoms would meet prespecified criteria in the L-CHSCY calibration and validation samples and would be supported in the available ONPATH age/informant groups.

## Methods

L-CHSCY. The 2023 Longitudinal Canadian Health Survey on Children and Youth (L-CHSCY) served as the primary dataset for this study. The L-CHSCY is a longitudinal follow-up survey conducted by Statistics Canada to examine the health and well-being of children, adolescents, and emerging adults across Canada. The survey follows participants originally recruited through the 2019 Canadian Health Survey on Children and Youth (CHSCY) and includes respondents residing in all ten Canadian provinces. Detailed information regarding the survey design, sampling procedures, coverage, weighting, response rates, and retention is available in the Statistics Canada User Guide (Statistics Canada. (2023) and L-CHSCY data resource profile paper (https://www23.statcan.gc.ca/imdb/p2SV.pl?Function=getSurvey&Id=1504253). Georgiades et al., 2026; *Preprint)*.

For the present analyses, participants were aged 5-21 years at follow-up and were grouped into three developmental stages: early and middle childhood (5-11 years; n = 11,860), adolescence (12-17 years; n = 7,240), and emerging adulthood (18-21 years; n = 3,640), yielding a total analytic sample of n =22,740. The person-most-knowledgeable (PMK) completed the survey on behalf of children aged 5-11 years, 98% identified as parents and so we will use the term parent’s hereafter. Adolescents and emerging adults aged 12–21 years self-completed the survey for most modules, including the OCHS-EBS item set.

ONPATH. The Ontario Provincial Assessment and Tracking of Child and Family Health Study (ONPATH) is a longitudinal population-based study of children and youth in Ontario. Participants were recruited through a probability-based online panel using stratified random sampling to approximate provincial demographic distributions. At baseline, 1,261 participants responded (29.7% response rate), and weighted characteristics aligned closely with provincial benchmarks. Detailed information regarding sampling procedures, weighting, response rates, and representativeness is available in the ONPATH methods paper and accompanying user guide (Georgiades et al., 2026 [pre-print]; Georgiades et al., 2026 [User Guide]).The present analyses used Wave 5 data collected from October to December 2022 (n=719 parent respondents and n=116 youth respondents). Parent reports were available for participants grouped as ages 4–11 and 12– 17 years, and youth self-reports were available for ages 12–17 years. Both informants completed internalizing and oppositional behaviour items; attention deficit/hyperactivity items were available in parent report only.

Age and sex assigned at birth were examined in both datasets. Sex assigned at birth was recorded as male or female and was used in subgroup and measurement invariance analyses.

Adapted-OCHS-EBS item set. Child and youth mental health symptoms were assessed using select items derived from the original 52-item OCHS-EBS ( Duncan et al., 2019). The items included in the present study were drawn from the Major Depressive Disorder (MDD), Generalized Anxiety Disorder (GAD), Oppositional Defiant Disorder (ODD), and Attention-Deficit/Hyperactivity Disorder (ADHD) domains of the original measure. Previous psychometric work using data from the 2014 Ontario Child Health Study identified an initial 29-item subset from these four domains for further evaluation. Subsequent exploratory factor analysis and item response theory analyses supported refinement to a 21-item version comprising eight internalizing items (MDD and GAD), six oppositional behaviour items, and seven attention deficit/hyperactivity items (see Georgiades et al., 2026, Appendix C [*pre-print*]). Consistent with this previous psychometric work, the primary analyses in the present study focused on this 21-item structure. Both the L-CHSCY and ONPATH datasets included these 21-items.

In the L-CHSCY, respondents reported symptoms during the previous six months; in ONPATH, respondents reported symptoms during the previous week. In both studies, items were scored 0 (never or not true), 1 (sometimes or somewhat true), or 2 (often or very true). Domain scores were calculated by summing responses to the retained items, with higher scores indicating more frequent symptoms.

### Analytic Approach

Pre-analysis checks were conducted without survey weights because the objective was to evaluate internal measurement structure rather than estimate population parameters (Furr, 2021). Item endorsement frequencies were examined within each age/informant group before factor analysis.

Factorial validity and item refinement. A random 50% L-CHSCY calibration sample spanning ages 5–21 years and both informants was used for exploratory factor analysis (EFA) of the 21 candidate items. Pooling was intentional: the aim was to identify a factor structure that could then be evaluated separately within each age/informant group. One-through five-factor models were estimated using weighted least squares mean- and variance-adjusted estimation with Geomin rotation. Model selection considered CFI, TLI, RMSEA, SRMR, rotated factor loadings, cross-loadings, and interpretability relative to the prespecified DSM-linked symptom domains.

The selected structure was evaluated by CFA in the remaining 50% L-CHSCY validation sample, overall and separately for ages 5–11 years (PMK report), 12–17 years (youth self-report), and 18– 21 years (self-report). Items were considered for removal using prespecified loading, cross-loading, model-fit, and convergent/discriminant-validity criteria together with consistency with the DSM-linked content specification.

Internal consistency reliability and validity evidence. Cronbach’s alpha was used to estimate internal consistency reliability. Convergent validity was evaluated using average variance extracted (AVE), with values above .50 meeting the prespecified criterion. Discriminant validity was supported when the square root of each factor’s AVE exceeded its correlations with other factors (Carter, 2016; Hair et al., 2021).

Measurement invariance. After the factor structure and internal consistency and validity criteria were evaluated, configural, metric, and scalar invariance across sex assigned at birth was examined separately within each L-CHSCY age/informant group. Cross-informant invariance was not tested because parent and youth reports were modelled separately due to only having one informant for each age group. The resulting L-CHSCY structure was then evaluated in ONPATH using CFAs for parent reports and youth self-reports, followed by the available within-informant invariance analyses, internal consistency, and validity.

## Results

### Psychometric Evaluation in the L-CHSCY

Factorial Validity and Item Refinement: EFA of the 21-item set in the L-CHSCY calibration sample (50% of the L-CHSCY dataset). Table 1 presents the fit indices for one-through five-factor solutions. Consistent with previous psychometric work, the EFA supported a three-factor structure corresponding to internalizing, oppositional behaviour, and inattention/hyperactivity symptoms. The three-factor solution demonstrated the most interpretable and theoretically coherent structure and was therefore retained for confirmatory analyses. Rotated item loadings are reported in Table 2.

**Table 1.** Fit indices for exploratory factor analyses of the 21-item candidate set in the L-CHSCY calibration sample (random 50%; ages 5–21 years)

| Model | X<sup>2</sup> | df | CFI | TLI | RMSEA | SRMR | EIG |
| --- | --- | --- | --- | --- | --- | --- | --- |
| 1- Factor | 25657.137* | 170 | 0.864 | 0.849 | 0.126 | 0.130 | 10.189 |
| 2-Factor | 12559.234 | 151 | 0.934 | 0.917 | 0.094 | 0.069 | 2.480 |
| 3-Factor | **3575.058\*** | **133** | **0.982** | **0.974** | **0.053** | **0.031** | **1.512** |
| 4-Factor | 1652.535* | 116 | 0.992 | 0.987 | 0.038 | 0.021 | 0.677 |
| 5-Factor | 1061.477* | 100 | 0.995 | 0.990 | 0.032 | 0.016 | 0.602 |

**Table 2.** Rotated item loadings for the three-factor exploratory factor analysis of the 20-item candidate set in the L-CHSCY calibration sample.

| Items (20 in total) | Factor 1<br>(OB) | Factor 2<br>(INT) | Factor 3<br>(ADHD) |
| --- | --- | --- | --- |
| A - Angry and resentful | **0.699** | 0.251 | -0.117 |
| B - Anxious or on edge | 0.001 | **0.896** | -0.041 |
| C - Argue a lot with adults | **0.768** | 0.005 | 0.012 |
| D - Blame others for mistakes | **0.713** | -0.123 | 0.064 |
| E - Can't concentrate/pay attention | -0.009 | 0.373 | **0.717** |
| F - Can't stay seated when required | 0.253 | -0.006 | **0.666** |
| I - Easily distracted | -0.012 | 0.352 | **0.732** |
| J - Easily annoyed by others | **0.404** | 0.412 | 0.064 |
| K - Fails to finish things you start | 0.155 | 0.297 | **0.511** |
| L - Feels worthless or inferior | 0.125 | **0.755** | 0.020 |
| M – Fidgets | 0.045 | 0.391 | **0.504** |
| N - Finds it hard to stop worrying | -0.106 | **0.916** | 0.029 |
| R - Has difficulty awaiting turn in games/groups | 0.482 | -0.088 | **0.394** |
| S - Have trouble enjoying self | 0.077 | **0.718** | 0.077 |
| T - Impulsive or acts without thinking | 0.505 | 0.096 | **0.385** |
| U - Loses temper | **0.800** | 0.062 | -0.014 |
| V - Nervous, high-strung or tense | -0.049 | **0.911** | 0.014 |
| W - Talks about killing self | 0.095 | **0.784** | -0.024 |
| X - Too fearful or anxious | -0.093 | **0.942** | -0.012 |
| Y - Unhappy, sad or depressed | 0.122 | **0.784** | -0.035 |

Note. Bold indicates the highest loading. ADHD = attention-deficit/hyperactivity disorder; EFA = exploratory factor analysis; INT = internalizing; L-CHSCY = Longitudinal Canadian Health Survey on Children and Youth; OB = oppositional behaviour.

Table 2 presents the rotated factor loadings from the EFA analysis on the calibration sample. Factor loadings indicated that the items loaded significantly on their respective factors. Based on factor loadings, these factors appear to represent internalizing, Oppositional Behaviour, and Attention-Deficit/Hyperactivity Disorder. The item “gets back at people” did not meet the prespecified loading and validity criteria in youth self-reports at ages 12-17 and 18-21 years, although it met the criteria in PMK reports at ages 5-11 years. Removing it improved youth model fit and discriminant validity. The item was removed for every informant to retain a common scoring set, yielding the 20-item OCHS-EBS-A: eight Internalizing, five Oppositional Behaviour, and seven Attention-Deficit/Hyperactivity Disorder items.

### Confirmatory Factor Analysis

The selected three-factor structure was evaluated in the independent L-CHSCY validation sample (remaining 50% of ample) and separately within each age/informant group. Fit statistics are reported in Table 3 and should be interpreted against the criteria specified in the Methods. The results supported the three-factor model with 20 items, demonstrating excellent fit indices across all age groups (CFI > 0.95, TLI > 0.90, RMSEA < 0.06, SRMR < 1).

**Table 3.** Confirmatory factor analysis fit indices of the three-factor, 20-item OCHS-EBS-A in the L-CHSCY validation sample, overall and by age/informant group.

| **Model** | **Age** | **X<sup>2</sup>** | **df** | **CFI** | **TLI** | **RMSEA** | **SRMR** |
| --- | --- | --- | --- | --- | --- | --- | --- |
| **3- Factor** | 5-21 | 4289.990* | 167 | 0.949 | 0.942 | 0.051 | 0.065 |
| **3- Factor** | 5-11 | 3977.112* | 187 | 0.957 | 0.952 | 0.059 | 0.064 |
| **3- Factor** | 12-17 | 1289.781* | 167 | 0.969 | 0.965 | 0.059 | 0.048 |
| **3- Factor** | 12-21 | 2692.386* | 187 | 0.970 | 0.966 | 0.049 | 0.053 |
| **3- Factor** | 18-21 | 1398.203* | 187 | 0.972 | 0.969 | 0.059 | 0.054 |

Two retained items had notable cross-loadings: “easily annoyed by others” across INT and OB, and “has difficulty awaiting turn in games/groups” across Oppositional Behaviour and Attention-Deficit/Hyperactivity Disorder. They were assigned to their prespecified DSM-linked factors because they met the retained-item criteria and the factor-level AVE and correlation results supported differentiation of the factors (Tables 4 and 5).

**Table 4.** Standardized factor loadings and factor correlations for the 20-item OCHS-EBS-A in the L-CHSCY validation sample.

| **Item** | **INT** | **OB** | **ADHD** |
| --- | --- | --- | --- |
| Anxious or on edge | 0.871 |  |  |
| Feels worthless or inferior | 0.842 |  |  |
| Finds it hard to stop worrying | 0.855 |  |  |
| Have trouble enjoying self | 0.800 |  |  |
| Nervous, high-strung or tense | 0.898 |  |  |
| Talks about killing self | 0.818 |  |  |
| Too fearful or anxious | 0.871 |  |  |
| Unhappy, sad or depressed | 0.835 |  |  |
| Angry and resentful |  | 0.768 |  |
| Argue a lot with adults |  | 0.701 |  |
| Blame others for mistakes |  | 0.569 |  |
| Easily annoyed by others |  | 0.840 |  |
| Loses temper |  | 0.770 |  |
| Can't concentrate/pay attention |  |  | 0.889 |
| Can't stay seated when required |  |  | 0.663 |
| Easily distracted |  |  | 0.863 |
| Fails to finish things you start |  |  | 0.762 |
| Fidgets |  |  | 0.739 |
| Has difficulty awaiting turn in games |  |  | 0.576 |
| Impulsive or acts without thinking |  |  | 0.769 |
| **Factors** | **OB** | **INT** | **ADHD** |
| **OB** | 1 |  |  |
| **INT** | 0.641 | 1 |  |
| **ADHD** | 0.717 | 0.666 | 1 |

**Table 5.** Convergent and discriminant validity evidence for OCHS-EBS-A scores by L-CHSCY age/informant group.

| Age |  | INT (8 items) | OB (5 items) | ADHD (7 items) |
| --- | --- | --- | --- | --- |
|  | **Convergent** | **AVE** | **AVE** | **AVE** |
| 5-11 | Passed | 0.658 | 0.749 | 0.653 |
| 12-17 | Passed | 0.717 | 0.587 | 0.528 |
| 12-21 | Passed | 0.727 | 0.622 | 0.556 |
| 18-21 | Passed | 0.731 | 0.629 | 0.571 |
|  | **Discriminant** | **SQRT AVE** | **SQRT AVE** | **SQRT AVE** |
| 5-11 | Passed | 0.811 | 0.866 | 0.808 |
| 12-17 | Passed | 0.847 | 0.766 | 0.727 |
| 12-21 | Passed | 0.852 | 0.789 | 0.746 |
| 18-21 | Passed | 0.855 | 0.793 | 0.756 |

**Note.** ADHD = attention-deficit/hyperactivity disorder; AVE = average variance extracted; INT = internalizing; L-CHSCY = Longitudinal Canadian Health Survey on Children and Youth; OB = oppositional behaviour; SQRT AVE = square root of average variance extracted.

Table 4 reports standardized CFA loadings and factor correlations. The values are presented for comparison with the prespecified thresholds.

### Internal Consistency Reliability and Validity Evidence

Cronbach’s alpha exceeded .80 for each factor within every L-CHSCY age/informant group (Appendix Table A1). These estimates provide evidence of internal consistency reliability only; temporal stability was not assessed.

AVE exceeded .50 for each factor within the reported age/informant groups, meeting the prespecified convergent-validity criterion. For each comparison, the square root of AVE exceeded the relevant inter-factor correlation, meeting the prespecified discriminant-validity criterion (Tables 4 and 5).

Within each L-CHSCY age/informant group, the configural, metric, and scalar invariance criteria across sex assigned at birth were met (Appendix Tables A2–A4). Parent and youth reports were analysed separately; cross-informant invariance was not tested.

### Psychometric Evaluation in ONPATH

In ONPATH, the L-CHSCY three-factor structure met the stated fit criteria in parent reports for ages 4–11 and 12–17 years. In youth self-report, the item “gets back at people” again did not meet the loading criteria. The issue was not observed in parent report. Removing this item improved model fit and discriminant validity between internalizing and Oppositional Behaviour.

After removing the item “gets back at people” from the OB factor, consistent with the refinement applied in the L-CHSCY, the model met criteria for discriminant validity for both informants.

Parent-report invariance findings are reported in the supplementary tables in appendix C1-C2.For ONPATH youth self-report, configural, metric, and scalar invariance criteria across sex assigned at birth were met for internalizing symptoms but not for OB symptoms, suggesting potential sex-based differences in how oppositional behaviours are interpreted or endorsed among youth.

## Discussion

In two independent population-representative samples, we evaluated an adapted OCHS-EBS item set (OCHS-EBS-A) across childhood, adolescence, and emerging adulthood. The L-CHSCY analysis identified a three-factor solution, and the validation analyses supported a refined 20-item OCHS-EBS-A representing Internalizing, Oppositional Behaviour, and Attention-Deficit/Hyperactivity Disorder symptoms. The reported scores met the study’s internal consistency and convergent and discriminant validity criteria within the available L-CHSCY for all three age groups and for parent and self-reports. ONPATH provided an additional evaluation rather than definitive replication. Parent-report findings supported the L-CHSCY structure meeting all the psychometric evaluation criteria for internal consistency, internal convergent and discriminant validity and measurement invariance. The youth self-report analysis reproduced the loading problem for the item “gets back at people.” Invariance criteria were met for youth-reported internalizing symptoms but not oppositional behaviour symptoms; the latter result should be interpreted cautiously due to the smaller youth sample.

Removing “gets back at people” improved youth model fit and discriminant validity in both studies. Although the item met criteria in parent report, a common 20-item set was retained across informants to preserve consistent scoring and implementation. This choice reflects a practical design decision previously used within the OCHS-EBS family; it does not imply that parent and youth reports should agree or that informant-specific versions could not perform differently. The observed cross-loadings are plausible as internalizing, oppositional behaviours, and inattention/hyperactivity symptoms overlap conceptually. Nonetheless, the retained items met the prespecified criteria, and factor-level AVE and correlation results supported differentiation. However, these analyses provide internal-structure evidence only; correlations with external constructs were not examined.

This is the first study to include a population-representative sample of children, adolescence and emerging adults across all ten provinces in Canada to evaluate the psychometric properties of items from the OCHS-EBS Additional evaluation in an independent sample speaks to the robustness of its psychometric properties.

Nevertheless, several study limitations deserve mention. Evidence of reliability was limited to evaluation of internal consistency. Test–retest reliability, inter-informant reliability, and sensitivity to change were not assessed. Second, Other limitations include the small ONPATH youth sample, absence of youth-reported inattention/hyperactivity items in ONPATH, different symptom-reference periods, lack of external construct-validity analyses, and underrepresentation of some population groups.

Despite these limitations, the OCHS-EBS-A (20-item version) reduces item count relative to the original 52-item OCHS-EBS and may lower administration time and respondent burden. These practical advantages were inferred from length, not directly evaluated; acceptability, completion time, and other aspects of utility require separate study. Future work should evaluate test–retest reliability, external construct validity, longitudinal invariance, and practical utility, and should assess the OCHS-EBS-A in more diverse populations.

The L-CHSCY analyses provide factorial-validity, internal-consistency, and convergent- and discriminant-validity evidence for scores from the 20-item OCHS-EBS-A among children, youth, and emerging adults. ONPATH provides additional, qualified evidence in a provincial sample. The results underscore the value of multi-informant approaches in population level child and youth mental health assessment. Discrepancies between youth and parent reports are common, particularly for externalizing behaviours (De Los Reyes & Kazdin, 2005). In our study, the problematic item was only evident in youth self-report and not in parent-report, suggesting that certain behavioural items may lack salience or clarity for adolescents. This finding raises important questions about whether items should be tailored for specific informants or whether a universal set of items can achieve measurement equivalence across reporters. Some scholars advocate for informant-specific item development, particularly when capturing behaviours that are situationally dependent or socially sensitive (Achenbach, 2011; Kraemer et al., 2003).

Further research is needed to explore how item design can better accommodate informant-specific nuances while maintaining the integrity of the scale. Although situated in Canada, these findings have broader international relevance for high-income countries. As countries increasingly implement national child health surveys, validated brief measures such as the OCHS-EBS-A are critical for ensuring comparability, monitoring trends, and informing coordinated responses to youth mental health.

Overall, this study sought to psychometrically evaluate an adapted version of the OCHS-EBS - the OCHS-EBS-A using data from a representative sample of children, youth, and emerging adults across all ten Canadian provinces (L-CHSCY), with additional validation using a provincial multi-informant dataset (ONPATH). Our findings underscore the broader significance of establishing reliable and valid measures of psychopathology for large epidemiologic population health studies among young people. Extrapolating from clinical samples cannot adequately represent the prevalence, course, or correlates of youth mental health problems, given that many affected individuals never present for services (Achenbach et al., 2005). Population-representative surveys such as the L-CHSCY therefore rely on robust measures to capture the scope of child and youth metal health symptoms across the population. Our evaluation demonstrates that the adapted 20-item OCHS-EBS-A is not only psychometrically sound, but also uniquely positioned as a scalable tool for epidemiologic surveys.

## Supporting information

Appendices OCHS EBS A

## Data Availability

Data from the Longitudinal Canadian Health Survey on CHildren and Youth (L-CHSCY) are available in Statistics Canada Research Data Centres and are not publicly available.
Data from the The Ontario Provincial Assessment and Tracking of Child and Family Health Study (ONPATH) are available upon reasonable request to the authors.

