## Appendices OCHS EBS A for "Psychometric Properties of a 20-Item Adapted Version of the Ontario Child Health Study Emotional Behavioural Scales for Population-Based Research"

#### Appendix 1 Measurement invariance for CHSCY results:

***Table A1.*** Internal Consistency values (Cronbach's alpha) for full 20-item scale and each distinct factor across age groups for L-CHSCY data

| **Age** | **INT (8 items)** | **OB (6 items)** | **ADHD (6 items)** | **ALL items** |
|---|---|---|---|---|
| **5-21 year olds.** | 0.997 | 0.996 | 0.996 | 0.999 |
| **5-11 year olds.** | 0.986 | 0.974 | 0.978 | 0.992 |
| **12-17 year olds.** | 0.999 | 0.998 | 0.998 | 0.999 |
| **18-21 year olds.** | 0.984 | 0.979 | 0.974 | 0.992 |

**Table A2** Measurement Invariance of CFA 3-factor for L-CHSCY data
Model by age 5-11 and by sex.

| | ***χ2*** | ***df*** | **p-value** | **RMSEA** | **CFI** | **SRMR** | **Decision** |
|---|---|---|---|---|---|---|---|
| **PMK report - Configural model** | 3842.400 | 334 | p<0.000 | 0.060 | 0.96 | 0.057 | **Passed** |
| **PMK report - Metric model** | 3845.280 | 351 | p<0.000 | 0.060 | 0.96 | 0.059 | **Passed** |
| **PMK report - Scalar model** | 3755.722 | 368 | p<0.000 | 0.056 | 0.96 | 0.057 | **Passed** |
| **PMK report -Configural vs. Metric** | ***Δχ2*** | ***Δdf*** | **p-value** | **ΔRMSEA** | **ΔCFI** | **ΔSRMR** | |
| | 56.204 | 17 | p<0.001 | 0 | 0 | -0.001 | |
| **PMK report -Configural vs. Scalar** | ***Δχ2*** | ***Δdf*** | **p-value** | **ΔRMSEA** | **ΔCFI** | **ΔSRMR** | |
| | 82.648 | 34 | p<0.001 | 0.004 | -0.001 | -0.001 | |
| **PMK report -Metric vs. Scalar** | ***Δχ2*** | ***Δdf*** | **p-value** | **ΔRMSEA** | **ΔCFI** | **ΔSRMR** | |
| | 31.898 | 17 | p<0.001 | 0.004 | -0.001 | 0 | |

**Table A3.** Measurement Invariance of CFA 3-factor for L-CHSCY data
Model by age 12-17 and by sex.

| | $\chi 2$ | *df* | **p-value** | **RMSEA** | **CFI** | **SRMR** | **Decision** |
|---|---|---|---|---|---|---|---|
| **YSR - Configural model** | 1326.716 | 334 | p<0.000 | 0.056 | 0.97 | 0.05 | **Passed** |
| **YSR - Metric model** | 1419.606 | 351 | p<0.000 | 0.057 | 0.97 | 0.051 | **Passed** |
| **YSR - Scalar model** | 1430.368 | 368 | p<0.000 | 0.055 | 0.97 | 0.052 | **Passed** |
| **YSR -Configural vs. Metric** | $\Delta\chi 2$ | *Δdf* | **p-value** | **ΔRMSEA** | **ΔCFI** | **ΔSRMR** | |
| | 104.283 | 17 | p<0.001 | -0.001 | 0.002 | -0.001 | |
| **YSR - Configural vs. Scalar** | $\Delta\chi 2$ | *Δdf* | **p-value** | **ΔRMSEA** | **ΔCFI** | **ΔSRMR** | |
| | 142.215 | 34 | p<0.001 | 0.001 | 0.002 | -0.002 | |
| **YSR-Metric vs. Scalar** | $\Delta\chi 2$ | *Δdf* | **p-value** | **ΔRMSEA** | **ΔCFI** | **ΔSRMR** | |
| | 56.537 | 17 | p<0.001 | 0.002 | 0 | -0.001 | |

**Table A4.** Measurement Invariance of CFA 3-factor for L-CHSCY data
Model by age 18-21 and by sex.

| | $\chi 2$ | *df* | **p-value** | **RMSEA** | **CFI** | **SRMR** | **Decision** |
|---|---|---|---|---|---|---|---|
| **YSR - Configural model** | 1606.902 | 334 | p<0.000 | 0.065 | 0.97 | 0.056 | **Passed** |
| **YSR - Metric model** | 1682.106 | 351 | p<0.000 | 0.065 | 0.97 | 0.056 | **Passed** |
| **YSR - Scalar model** | 1654.474 | 368 | p<0.000 | 0.062 | 0.97 | 0.057 | **Passed** |
| **YSR - Configural vs. Metric** | $\Delta\chi 2$ | *Δdf* | **p-value** | **ΔRMSEA** | **ΔCFI** | **ΔSRMR** | |
| | 91.996 | 17 | p<0.001 | 0 | 0.002 | 0 | |
| **YSR - Configural vs. Scalar** | $\Delta\chi 2$ | *Δdf* | **p-value** | **ΔRMSEA** | **ΔCFI** | **ΔSRMR** | |
| | 111.044 | 34 | p<0.001 | 0.003 | 0 | -0.001 | |
| **YSR - Metric vs. Scalar** | $\Delta\chi 2$ | *Δdf* | **p-value** | **ΔRMSEA** | **ΔCFI** | **ΔSRMR** | |
| | 34.815 | 17 | p<0.001 | 0.003 | -0.002 | -0.001 | |

Note: YSR= youth self report.

Assessments of configural, metric and scalar models all passed indicating the 3-factor 20 item model is consistent and reliable across these three measurement invariance assessments.

### Appendix 2: ONPATH results

Exploratory Factor Analysis (EFA) was conducted on parent-reported data in the ONPATH sample, identifying a three-factor structure consistent with the domains of Internalizing, Oppositional Behaviours (OB), and ADHD. Given the focus on informant discrepancies observed in the CHSCY dataset, follow-up Confirmatory Factor Analyses (CFA) were conducted separately for parent reports (ages 4–17) and youth self-reports (ages 12–17), with a focus on the Internalizing and OB domains.

#### Parent Reports

CFA models for parent-reported data showed acceptable model fit across the full sample (ages 4–17), as well as within age subgroups (4–11 and 12–17) (Appendix 2 Tables 7a-7c). No issues were identified with the item *"gets back at people"*, contrasting with the patterns observed in youth self-report data.

#### Youth Self-Reports (Ages 12–17)

In line with CHSCY findings, CFA results for youth self-reports indicated poor factor loadings and cross-loadings for the item *"gets back at people"*, which negatively affected model fit (Appendix 2 Table 8). After removing the item, model fit improved substantially, validating the refined 20-item model for youth self-report.

#### Measurement Invariance Testing

Measurement invariance testing across sex in the ONPATH youth self-report data yielded mixed results. The Internalizing domain demonstrated configural, metric, and scalar invariance, indicating that the factor structure and item functioning were equivalent across male and female adolescents. However, the Oppositional Behaviour domain did not meet criteria for invariance, suggesting that these items may be interpreted or endorsed differently across sex (Appendix 2 Table 9). This pattern is consistent with previous research highlighting sex-based differences in the self-reporting of externalizing behaviours during adolescence. Rather than reflecting a failure of the scale, these findings underscore the importance of testing invariance at the domain level and cautioning against direct sex-based comparisons in youth-reported OB symptoms. Future work should explore item-level non-invariance and assess the feasibility of establishing partial invariance for more meaningful group comparisons.

**Table B1** CFA Results for Parent Reports ONPATH (Ages 5–17) for Internalizing, OB, and ADHD for ONPATH data

| **Factor** | **Item** | **Loading** | **$R^2$** |
|---|---|---|---|
| **Internalizing** | Anxious or on edge | 0.894 | 0.80 |
| | Feels worthless or inferior | 0.894 | 0.71 |
| | Finds it hard to stop worrying | 0.870 | 0.76 |
| | Has trouble enjoying self | 0.754 | 0.57 |
| | Nervous, high-strung or tense | 0.890 | 0.79 |
| | Talks about killing self | 0.801 | 0.64 |
| | Fearful or anxious | 0.886 | 0.78 |
| | Unhappy, sad or depressed | 0.787 | 0.62 |
| Internalizing AVE | - | - | 0.692 |
| Internalizing √AVE | - | - | 0.832 |
| Discriminant Validity | | | **Passed** |
| **ADHD** | Can't concentrate, can't pay attention for long | 0.884 | 0.78 |
| | Can't stay seated when required to do so | 0.764 | 0.58 |
| | Distractible, has trouble sticking to any activity | 0.876 | 0.77 |
| | Fails to finish things he/she starts | 0.742 | 0.55 |
| | Fidgets | 0.738 | 0.54 |
| | Has difficulty awaiting turn in games or groups | 0.593 | 0.35 |
| | Impulsive or acts without thinking | 0.766 | 0.59 |
| ADHD AVE | - | - | 0.595 |
| ADHD √AVE | - | - | 0.771 |
| Discriminant Validity | - | | **Passed** |
| **Oppositional Behaviour** | Angry and resentful | 0.869 | 0.76 |
| | Argues a lot with adults | 0.756 | 0.57 |
| | Blames others for own mistakes | 0.666 | 0.44 |
| | Easily annoyed by others | 0.787 | 0.62 |
| | Gets back at people | 0.536 | 0.29 |
| | Loses temper | 0.810 | 0.66 |
| Oppositional AVE | - | - | 0.556 |
| Oppositional √AVE | - | - | 0.745 |
| Discriminant Validity | | | **Passed** |
| **Factor Correlations** | **OB** | **INT** | |
| ADHD | 0.669 | 0.53 | - |
| INT | 0.702 | - | - |

**Table B2** CFA Results for Parent Reports ONPATH (Ages 5–11) for Internalizing, OB, and ADHD for ONPATH data

| **Factor** | **Item** | **Loading** | **$R^2$** |
|---|---|---|---|
| **Internalizing** | Anxious or on edge | 0.915 | 0.84 |
| | Feels worthless or inferior | 0.815 | 0.66 |
| | Finds it hard to stop worrying | 0.867 | 0.75 |
| | Has trouble enjoying self | 0.758 | 0.57 |
| | Nervous, high-strung or tense | 0.913 | 0.83 |
| | Talks about killing self | 0.533 | 0.28 |
| | Fearful or anxious | 0.839 | 0.70 |
| | Unhappy, sad or depressed | 0.715 | 0.51 |
| Internalizing AVE | - | - | 0.645 |
| Internalizing √AVE | - | - | 0.803 |
| Discriminant Validity | | | **Passed** |
| **ADHD** | Can't concentrate, can't pay attention for long | 0.863 | 0.74 |
| | Can't stay seated when required to do so | 0.818 | 0.67 |
| | Distractible, has trouble sticking to any activity | 0.822 | 0.68 |
| | Fails to finish things he/she starts | 0.691 | 0.48 |
| | Fidgets | 0.757 | 0.57 |
| | Has difficulty awaiting turn in games or groups | 0.599 | 0.36 |
| | Impulsive or acts without thinking | 0.739 | 0.55 |
| ADHD AVE | - | - | 0.578 |
| ADHD √AVE | - | - | 0.760 |
| Discriminant Validity | - | - | **Passed** |
| **Oppositional Behavior** | Angry and resentful | 0.876 | 0.77 |
| | Argues a lot with adults | 0.739 | 0.55 |
| | Blames others for own mistakes | 0.596 | 0.36 |
| | Easily annoyed by others | 0.779 | 0.61 |
| | Gets back at people | 0.624 | 0.39 |
| | Loses temper | 0.793 | 0.63 |
| Oppositional AVE | - | - | 0.549 |
| Oppositional √AVE | - | - | 0.741 |
| Discriminant Validity | - | - | **Passed** |
| **Factor Correlations** | **OB** | **INT** | |
| INT | 0.651 | 0.502 | - |
| ADHD | 0.695 | - | - |

**Table B3.** CFA Results for Parent Reports ONPATH (Ages 12–17) for Internalizing, OB, and ADHD for ONPATH data

| Factor | Item | Loading | $R^2$ |
|---|---|---|---|
| **Internalizing** | Anxious or on edge | 0.885 | 0.78 |
| | Feels worthless or inferior | 0.857 | 0.73 |
| | Finds it hard to stop worrying | 0.868 | 0.75 |
| | Has trouble enjoying self | 0.757 | 0.57 |
| | Nervous, high-strung or tense | 0.862 | 0.74 |
| | Talks about killing self | 0.871 | 0.76 |
| | Fearful or anxious | 0.932 | 0.87 |
| | Unhappy, sad or depressed | 0.842 | 0.71 |
| Internalizing AVE | - | - | 0.740 |
| Internalizing √AVE | - | - | 0.860 |
| Discriminant Validity | | | **Passed** |
| **ADHD** | Can't concentrate, can't pay attention for long | 0.895 | 0.80 |
| | Can't stay seated when required to do so | 0.73 | 0.53 |
| | Distractible, has trouble sticking to any activity | 0.913 | 0.83 |
| | Fails to finish things he/she starts | 0.805 | 0.65 |
| | Fidgets | 0.712 | 0.51 |
| | Has difficulty awaiting turn in games or groups | 0.624 | 0.39 |
| | Impulsive or acts without thinking | 0.796 | 0.63 |
| ADHD AVE | - | - | 0.621 |
| ADHD √AVE | - | - | 0.788 |
| Discriminant Validity | - | - | **Passed** |
| **Oppositional Behavior** | Angry and resentful | 0.881 | 0.78 |
| | Argues a lot with adults | 0.785 | 0.62 |
| | Blames others for own mistakes | 0.757 | 0.57 |
| | Easily annoyed by others | 0.83 | 0.69 |
| | Gets back at people | 0.459 | 0.21 |
| | Loses temper | 0.856 | 0.73 |
| Oppositional AVE | - | - | 0.600 |
| Oppositional √AVE | - | - | 0.774 |
| Discriminant Validity | - | - | **Passed** |
| **Factor Correlations** | **OB** | **INT** | |
| INT | 0.704 | 0.648 | - |
| ADHD | 0.708 | - | - |

**Table B4** CFA Results for Youth Self-Reports ONPATH (Ages 12–17) for Internalizing and OB for ONPATH data

| Factor | Item | Loading | $R^2$ |
|---|---|---|---|
| INT | I feel worthless or inferior | 0.835 | 0.70 |
| | I have trouble enjoying myself | 0.854 | 0.73 |
| | I think about killing myself | 0.957 | 0.92 |
| | I am unhappy, sad or depressed | 0.835 | 0.70 |
| | I am anxious or on edge | 0.771 | 0.59 |
| | I find it hard to stop worrying | 0.73 | 0.53 |
| | I am nervous or tense | 0.829 | 0.69 |
| | I am fearful or anxious | 0.906 | 0.82 |
| Internalizing AVE | - | - | 0.709 |
| Internalizing √AVE | - | - | 0.842 |
| Discriminant Validity | - | - | **Passed** |
| OB | I am angry and resentful | 0.92 | 0.85 |
| | I argue a lot with adults | 0.622 | 0.39 |
| | I blame others for my own mistakes | 0.582 | 0.34 |
| | I am easily annoyed by others | 0.707 | 0.50 |
| | I lose my temper | 0.718 | 0.52 |
| Oppositional AVE | - | - | 0.517 |
| Oppositional √AVE | - | - | 0.719 |
| Discriminant Validity | - | - | **Passed** |
| **Factor Correlations** | **OB** | - | - |
| **INT** | 0.719 | - | - |

**Note:** Discriminant validity only passed when the item "Gets back at people" was removed.

**Table C1Parent report** - Testing Measurement Invariance of Confirmatory Factor Analysis of 3 factors **by sex** for ONPATH data

| | ***χ2*** | ***df*** | **p-value** | **RMSEA** | **CFI** | **SRMR** | **Decision** |
|---|---|---|---|---|---|---|---|
| PMK report - Configural model | 764.953 | 354 | p<0.001 | 0.043 | 0.93 | 0.058 | **Passed** |
| PMK report - Metric model | 775.886 | 372 | p<0.001 | 0.042 | 0.93 | 0.061 | **Passed** |
| PMK report - Scalar model | 821.336 | 390 | p<0.001 | 0.042 | 0.93 | 0.063 | **Passed** |
| **PMK report -Configural vs. Metric** | ***Δχ2*** | ***Δdf*** | **p-value** | **ΔRMSWA** | **ΔCFI** | **ΔSRMR** | |
| | 15.993 | 18 | p=0.593 | 0.001 | -0.001 | -0.003 | |
| **PMK report -Configural vs. Scalar** | ***Δχ2*** | ***Δdf*** | **p-value** | **ΔRMSWA** | **ΔCFI** | **ΔSRMR** | |
| | 58.599 | 36 | p=0.01 | 0.001 | 0.003 | -0.005 | |
| **PMK report -Metric vs. Scalar** | ***Δχ2*** | ***Δdf*** | **p-value** | **ΔRMSWA** | **ΔCFI** | **ΔSRMR** | |
| | 45.357 | 18 | p<0.001 | 0 | 0.004 | -0.002 | |

**Table C2 Parent report**- Testing Measurement Invariance of Confirmatory Factor Analysis of 3 factors **by age** For ONPATH data

| | ***χ2*** | ***df*** | **p-value** | **RMSEA** | **CFI** | **SRMR** | **Decision** |
|---|---|---|---|---|---|---|---|
| PMK report - Configural model | 778.538 | 354 | p<0.001 | 0.044 | 0.93 | 0.056 | **Passed** |
| PMK report - Metric model | 858.326 | 372 | p<0.001 | 0.046 | 0.92 | 0.067 | **Passed** |
| PMK report - Scalar model | 997.928 | 390 | p<0.001 | 0.05 | 0.91 | 0.069 | **Passed** |
| **PMK report -Configural vs. Metric** | ***Δχ2*** | ***Δdf*** | **p-value** | **ΔRMSWA** | **ΔCFI** | **ΔSRMR** | |
| | 74.748 | 18 | p<0.001 | -0.002 | 0.01 | -0.011 | |
| **PMK report -Configural vs. Scalar** | ***Δχ2*** | ***Δdf*** | **p-value** | **-0.006** | **ΔCFI** | **ΔSRMR** | |
| | 207.913 | 36 | p<0.001 | -0.006 | 0.022 | -0.013 | |
| **PMK report -Metric vs. Scalar** | ***Δχ2*** | ***Δdf*** | **p-value** | **ΔRMSWA** | **ΔCFI** | **ΔSRMR** | |
| | 136.101 | 18 | p<0.001 | -0.004 | 0.012 | -0.002 | |

**Note:** Measurement invariance testing was conducted within informant groups and tailored to the structure of the available data. For parent reports, invariance was evaluated across age groups (4–11 and 12–17) and by sex, as parent data spanned multiple developmental stages. In contrast, youth self-report data were available only for adolescents aged 12–17, precluding age-based invariance testing. Accordingly, invariance in youth data was assessed by sex only. Given domain-specific concerns identified in preliminary analyses, measurement invariance in youth self-report was examined separately for the Internalizing and Oppositional Behaviour domains.

**Table C3 Youth report** - Testing Measurement Invariance of Confirmatory Factor Analysis of OB **by sex** for ONPATH data

| | *χ2* | *df* | **p-value** | **RMSEA** | **CFI** | **SRMR** | **Decision** |
|---|---|---|---|---|---|---|---|
| YSR - Configural model | 283.434 | 134 | p<0.001 | 0.09 | 0.858 | 0.091 | **failed** |
| YSR - Metric model | 314.135 | 146 | p<0.001 | 0.101 | 0.841 | 0.115 | **failed** |
| YSR - Scalar model | 339.828 | 158 | p<0.001 | 0.1 | 0.828 | 0.118 | **failed** |
| **YSR - Configural vs. Metric** | *Δχ2* | *Δdf* | **p-value** | **ΔRMSWA** | **ΔCFI** | **ΔSRMR** | |
| | 29.171 | 12 | p<0.001 | -0.011 | 0.017 | -0.024 | |
| **YSR - Configural vs. Scalar** | *Δχ2* | *Δdf* | **p-value** | **-0.01** | **ΔCFI** | **ΔSRMR** | |
| | 54.97 | 24 | p<0.001 | -0.01 | 0.03 | -0.027 | |
| **YSR - Metric vs. Scalar** | *Δχ2* | *Δdf* | **p-value** | **ΔRMSWA** | **ΔCFI** | **ΔSRMR** | |
| | 25.723 | 12 | p<0.001 | 0.001 | 0.013 | -0.003 | |

*Note: YSR = youth self report. ADHD domain was not included in youth self-report data and therefore excluded from youth analyses.*

**Table C4 Youth report** - Testing Measurement Invariance of Confirmatory Factor Analysis of **INT by sex** for ONPATH data

| | *χ2* | *df* | **p-value** | **RMSEA** | **CFI** | **SRMR** | **Decision** |
|---|---|---|---|---|---|---|---|
| YSR - Configural model | 73.542 | 28 | p<0.001 | 0.119 | 0.931 | 0.071 | **Passed** |
| YSR - Metric model | 83.686 | 35 | p<0.001 | 0.11 | 0.936 | 0.107 | **Passed** |
| YSR - Scalar model | 93.928 | 42 | p<0.001 | 0.104 | 0.921 | 0.109 | **Passed** |
| **YSR - Configural vs. Metric** | *Δχ2* | *Δdf* | **p-value** | **ΔRMSWA** | **ΔCFI** | **ΔSRMR** | |
| | 12.868 | 7 | 0.0754 | 0.009 | -0.005 | -0.036 | |
| **YSR - Configural vs. Scalar** | *Δχ2* | *Δdf* | **p-value** | **0.015** | **ΔCFI** | **ΔSRMR** | |
| | 24.388 | 14 | 0.0411 | 0.015 | 0.01 | -0.038 | |
| **YSR - Metric vs. Scalar** | *Δχ2* | *Δdf* | **p-value** | **ΔRMSWA** | **ΔCFI** | **ΔSRMR** | |
| | 11.403 | 7 | 0.1220 | 0.006 | 0.015 | -0.002 | |

*Note: YSR= youth self report. ADHD domain was not included in youth self-report data and therefore excluded from youth analyses.*
